# Caring for Terminally Ill Cancer Patients: A Systematic Review of Literature on the Stress Experienced by Family Caregivers

**DOI:** 10.1101/2023.09.20.23295878

**Authors:** Kaijyun Jhang, Dih-Ling Luh

**Author notes:** Correspondence: **Dih-Ling Luh, E-mail:**.

## Abstract

**Objective:** This research takes previous study, Cancer family caregivers during the palliative, hospice, and bereavement phases: A review of the descriptive psychosocial literature, limited in recent decade, as methodology template. The purpose of this review was to organize the literature as compared to the different result of previous study.

**Method:** As a systematic review, major databases were searched for non-intervention descriptive studies. Psychosocial variables of family caregivers to adults with cancer during the different phases would be included.

**Result:** The 23 studies reviewed were conducted in ten countries and varied considerably by samples, outcome measures, and results. Despite limiting several conditions, results, such as age, gender, and relationship to the patient, were inconsistent. Across the 23 studies, 53 unique instruments were used; 13 of which were no psychometric testing. The family caregivers who were younger and faced level of daily life impairment tended to be burden, anxious, depress. To summarize the different factors influencing caregivers’ status, complicated grief was consistent with their situation.

**Conclusion:** As comparewith previous study, it demonstrated inconsistent results, which were spouse, gender and age, affecting family caregivers’ status. However, regarding to measurement instruments using, it was much more rigorous than before. Also, it had been changed in the major study site and the number of study. As a consequence of physical and psychosocial status of family caregivers, they were in high risk population.

## INTRODUCTION

### Hospice service

Hospice and palliative care system nowadays was originated from the United Kingdom by 1967(Xie Wei Ming, 2014). At the beginning of the twentieth century, this type of caring system was introduced to Taiwan; after that, it made medical community realize that severe patients, such as cancer or major organ failure, was better to let them feel comfortable and respectable than living in deteriorating but waiting to die alone (Li Chia Yu, 2010). The previous research discussed (Merydawilda Colón, 2014) indicated that various factors deeply affected Latino utilization of hospice care. These factors include beliefs about health care, death and end-of-life care, lack of insurance, lower referral rates by health care professionals and the hospice caregiver requirement. Government in Taiwan has actively performed and propagated hospice caring in home and community (Hospice Palliative Care Act). It should make patients not only under medical care but also surrounded by their family; if patients would like to receive spiritual care, there is certificated religious prayer doing benediction so as to show confident on returning back to Elysium. While patients departed from this life, reminiscence between patients and their family portrayed a panorama of ups and downs. It also made family relieved. The death of patient is a question of medical failure; but if it failed to assist patients to die in dignity, it is definitely a medical failure. Therefore, to maintain patient’s quality of life instead of ineffective medical care is prior discussion in the future.

### Family caregiver

After all these years, topic issue was always focused on welfare of dementia and physical and mental disabilities, but it ignored the family caregiver, who was being in shadow and silence (CY Hsiao, 2010). While caring for patients, family caregivers took them as priority; there is no purpose on their life without patients. They pay no regret on their loved one. However, as long as patients passed away, because of physically and mentally exhausted, no savings and difficult to return to workplace, the family caregiver possibly became the poor population. In the face of aging population, it was gradually being followed with interest on family caregiver in the developed country.

Family caregiver paid an important role on long term caring system, so it should render particularly on policy support; it implicated that caregiver can extend their duration of caring but also prevent their health status from stress or other illness because of caring. If family caregiver’s health status had deteriorated, patient’s quality of life decreased as well; on the other world, they would be forced to send to nursing home by necessary. Thus, it is a major issue to provide a supportive service for family caregiver in the purpose of reducing burden on caring. So, we shall create an amiable environment for patient and family caregiver at home place.

### Hospice

“Patient Right to Autonomy Act”, published at January 6, 2016, was stipulated to respect patient autonomy in healthcare, to safeguard their rights to a good death, and to promote a harmonious physician-patient-relationship. A patient, who has made an advance decision, suffered from terminally ill, irreversible coma, permanent vegetative state and severe dementia, then the medical institution or physician may, in accordance with the advance decision, partially or fully terminate, withdraw, or withhold life-sustaining treatments, artificial nutrition and hydration (Ministry of Justice, ROC). End-of-life implicated that many kinds of aspects towards death and expectation in the society (Hu Wunyu, 2005). Definitions of end-of-life was diverse in different culture and times; then it took a leap forward to adapt diversity in the final chapter. If there was a way to satisfy the expectation of patients and families, it can, in accord with clinical, cultural and ethical standards, eliminate the frustration and misfortune of patients, families, and caregivers. Thus, the end of life may be competed (Institute of Medicine National Academy of Science, 1997)

### Bereavement

Bereavement was a slow process from birth to death and death to be alive, which made people experience different step on grief. People in bereaved would suddenly be anger, deniable, bargaining or even depress (Deborah P. Waldrop, 2008). After psychological mechanism, their paralyzed thought would gradually fade away; It might take into healed and balance instead of desperate feeling on significant loss.

People undergone this situation possibly lasting less than few months or even being several years. Sadness and despair tool place repeating again and again; grief, fear, hopeless, imbalance, solitude and heartbroken feeling often strike quietly that makes people departed from normal life (Grande, Gunn E, 2004). If grief hadn’t been got appropriate catharsis, it might become eternal melancholy (Margaret Stroebe, 2008). It was best to face positively and actively on bereavement; so long as we came in front of it, sorrowful experience would be disappeared. Treatment of trauma probably took one or two years; but if we accepted and honestly confront sorrowful feeling, pain shall be end (TA Rando, 1993). In the process of being death to alive, people found their way continuously on life and attempt to stand at a point among many aspects of impact. Living and death definitely can form a profound but unique experience and feeling allowing particular training and gift for all of us.

We apply a previous research, Cancer family caregivers during the palliative, hospice, and bereavement phases: A review of the descriptive psychosocial literature (Anna-Leila Williams, 2010), as reference. Because a caregiver’s perceived burden and psychosocial concerns might be different at different phases of the patient’s disease, it brought them physically and mentally exhausted. Usually, the psychological burden of the caregiver was underestimated as compared with the stress of the terminal ill patient. The precious review urgently suggested to develop research standards, especially regarding measurement instruments, so it could intervene objectively and support actively family caregiver. Thus, and so, this literature review tended to find out the research on the family caregivers of terminal ill patients in the last ten years. With different phase of challenge, it should figure out that the truth and contradiction by experience and research result.

## METHOD

At first, we calibrated review article “Cancer family caregivers during the palliative, hospice, and bereavement phases: A review of the descriptive psychosocial literature” (Anna-Leila Williams, 2011) as referral formwork. The major database was searched in PubMed from 2008∼2016. The terms “caregiver,” “caregiving,” “neoplasm,” “oncology,” and “cancer” were entered as keywords. In addition, circumscription of website program was set in “’caregiver’ OR ‘caregiving’” AND “’neoplasm’ OR ‘oncology’ OR ‘cancer’”

Results were limited to English language, non-intervention descriptive studies, and sample was included from psychosocial variables of family caregivers to adult patients. As for results by year were shown in figure 1502 articles were found; afterwards, it was selected artificially out in 23 articles. This study had finished looking for the research article at the beginning of 2017, so the research after the beginning of 2017 was excluded (Figure 1).

**Figure 1.**
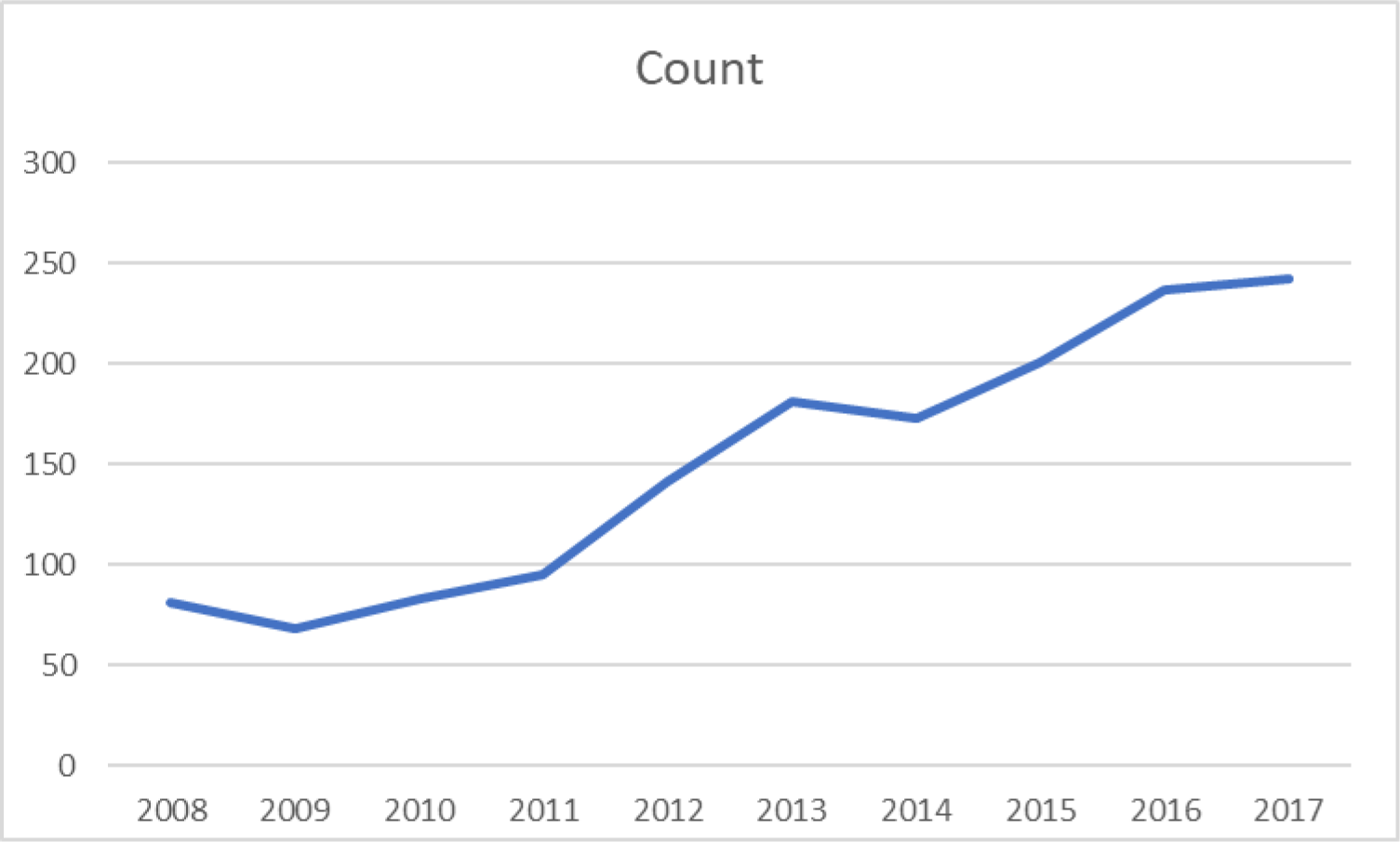
The number of publications from 2000 to 2017.

## RESULT

Despite limiting the articles reviewed to descriptive studies of cancer family caregivers in the palliative, hospice, or post-death bereavement phases, there remained considerable heterogeneity relative to caregiver characteristics, patient characteristics, measurement tools, and analytic methods. The 23 English language studies were conducted in 10 countries: Korea (7 studies), USA (7 studies), Taiwan (3 studies), Germany (1 studies), France (1 studies), Denmark (1 studies), Austria (1 studies), Canada (1 studies), Indonesia (1 studies), United Kingdom (1 studies). Study sample sizes ranged from 56 to 3560. Of the 23 studies, 13% (3studies) had a sample size of <100 family caregivers.

Three studies of the 23 were longitudinal in design, and 14 of which were cross sectional study; however, two of the studies (Yong Joo Lee, 2015; Young Sun Rhee, 2008) used similar material and method. Those of two cross-sectional studies show several detriments influencing family caregiver’ s quality of life which burdensomeness, younger age, disruptiveness and low social support or income were associated with emotional distress.

Most of the studies were of caregivers who attended to patients with a variety of cancers; only two studies focused on caregivers to individuals with a particular type of cancer: head and neck cancer (Chandylen L Nightingale, 2016), gastrointestinal cancer and lung cancer (Jamie M. Jacobs/2017). The two studies that looked at a particular type of cancer each had, respectively, a sample size of 56 and 275 participants. The small number samples made it difficult to interpret the influence of cancer type on the caregiver experience; relatively, enough quantity of sample made confidence in study research.

The extant literature regularly ascribed the relationship of the caregiver to the care recipient (spouse/partner, adult child, friend and so on) as a major influence on the caregiving experience. Of the 23 studies sample to gender, exclusively. One study focused on age.Three studies did not report the caregiver’ s relationship to the care recipient, and the remaining 20 studies had caregiver samples that were composed of individuals with a variety of different relationships with the care recipient.

Besides, 8 studies compared spouses versus non-spouses on psychosocial outcomes. Results were inconsistent. When comparing spouses versus non-spouses in the palliative/hospice phase, Tanguy Leroy (2015), Jamie M. Jacobs (2017) and Yong Joo Lee (2015) found no significant difference between groups. Paradoxically, research from Young Sun Rhee (2008) and Debra Parker Oliver (2016) revealed worse psychosocial outcomes during hospice/palliative phase as compared to Mette Kjaergaard Nielsen (2016) and Christantie Effendy (2015) report non-spouses that had worse psychosocial outcomes. Two study in the bereaved phase that compared spouses versus non-spouses found negative influence from the relationship (H. Götze, 2016; Sing-Fang Ling, 2013) (Table 2).

**Table 1.**
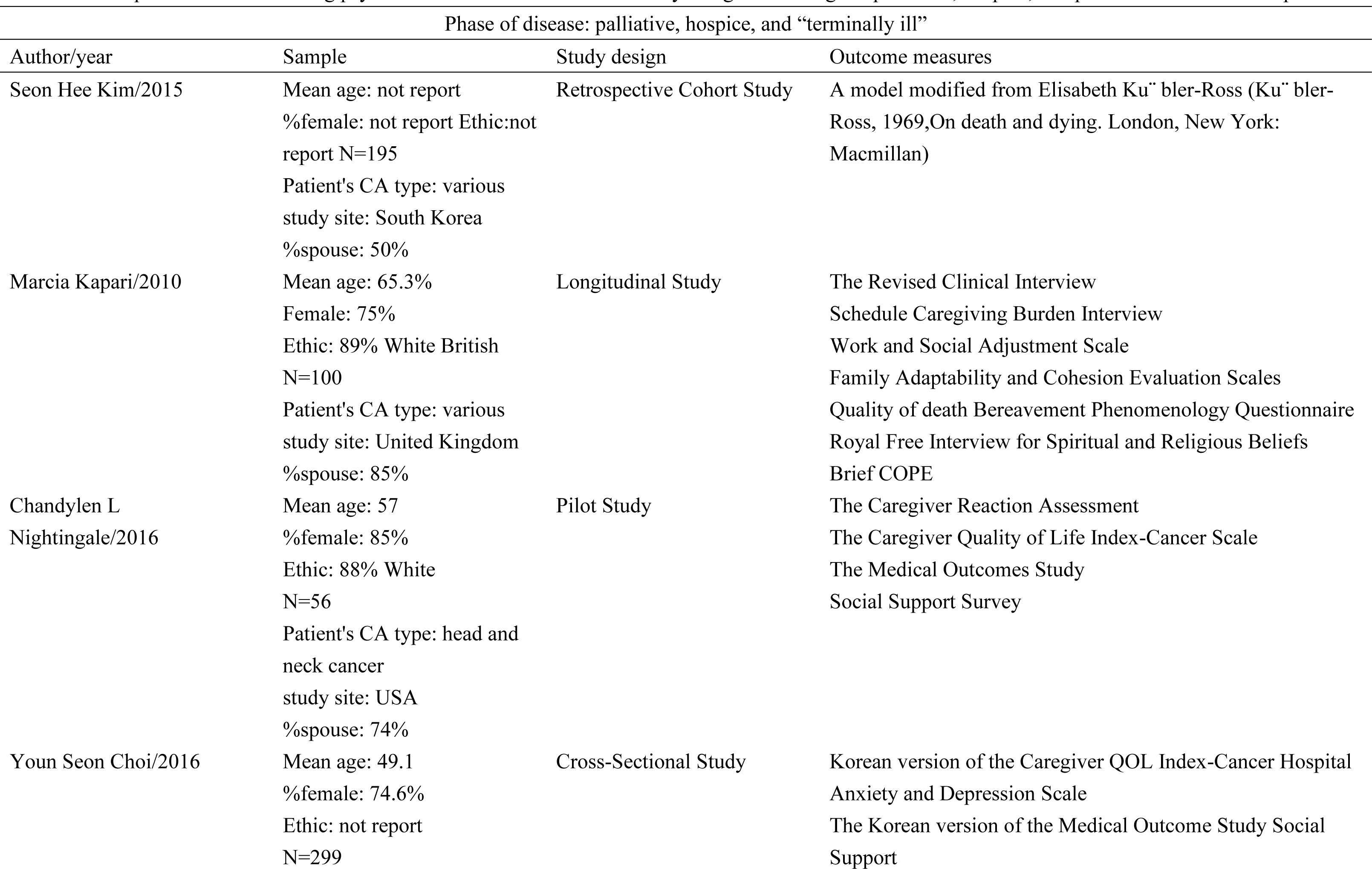
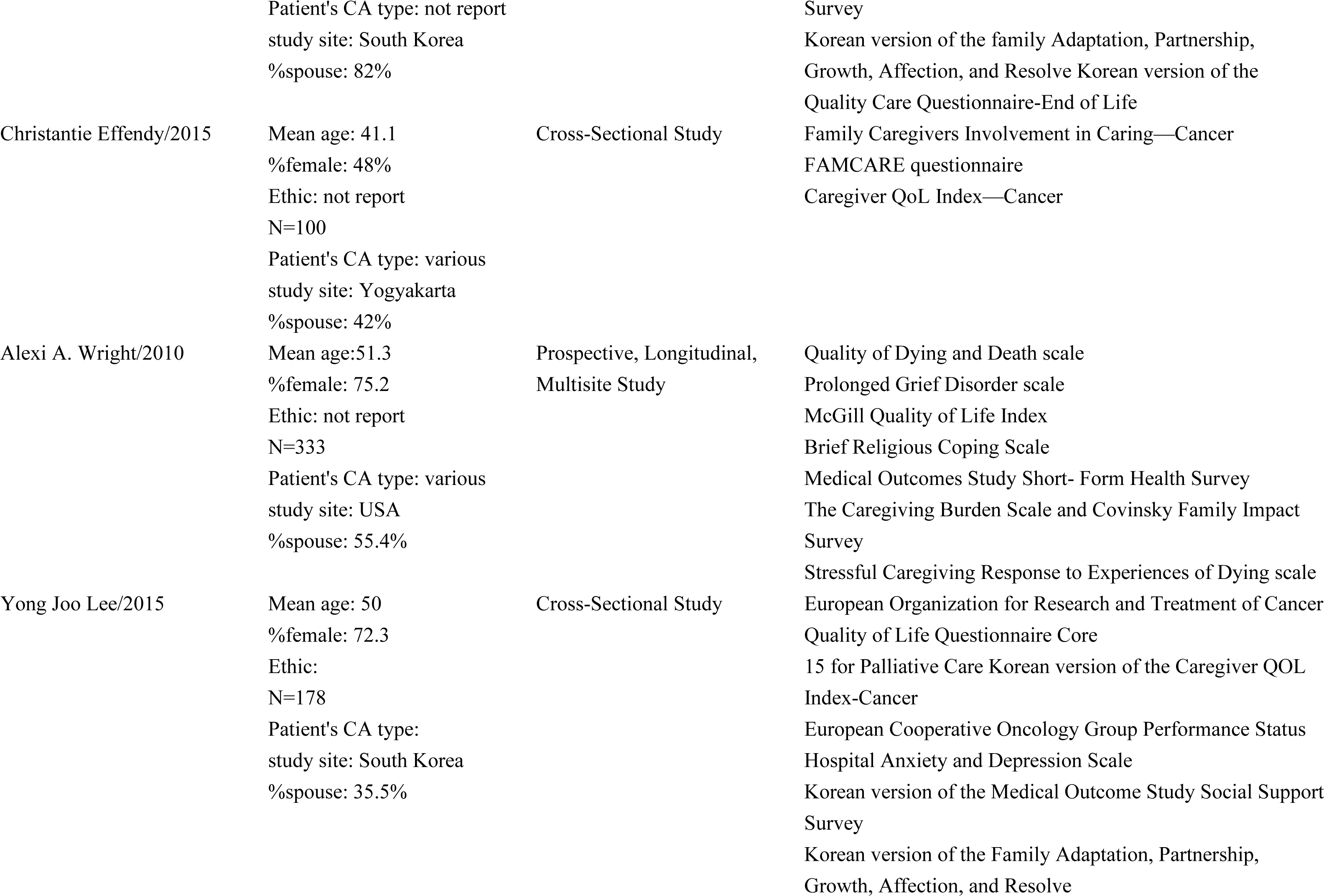
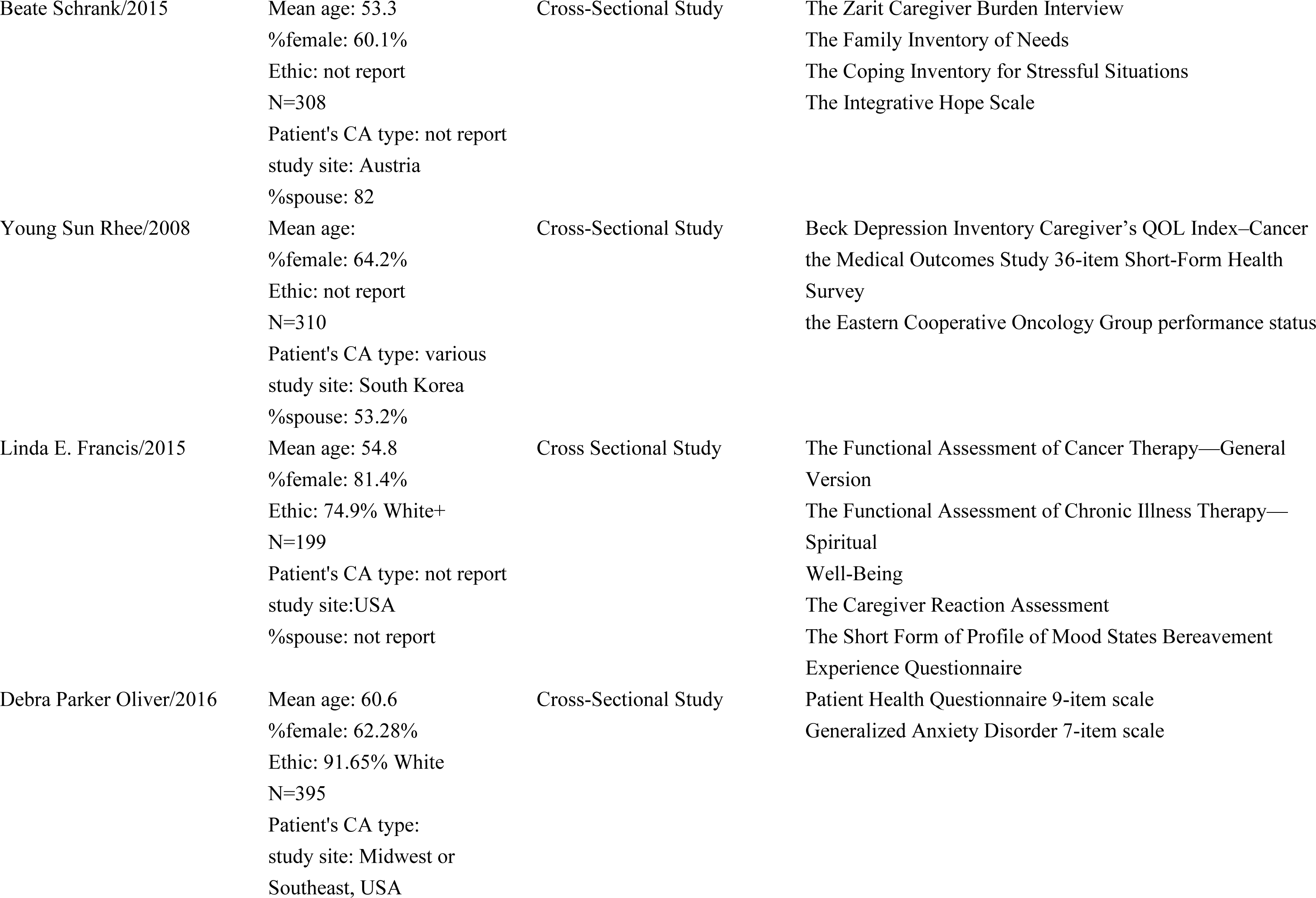
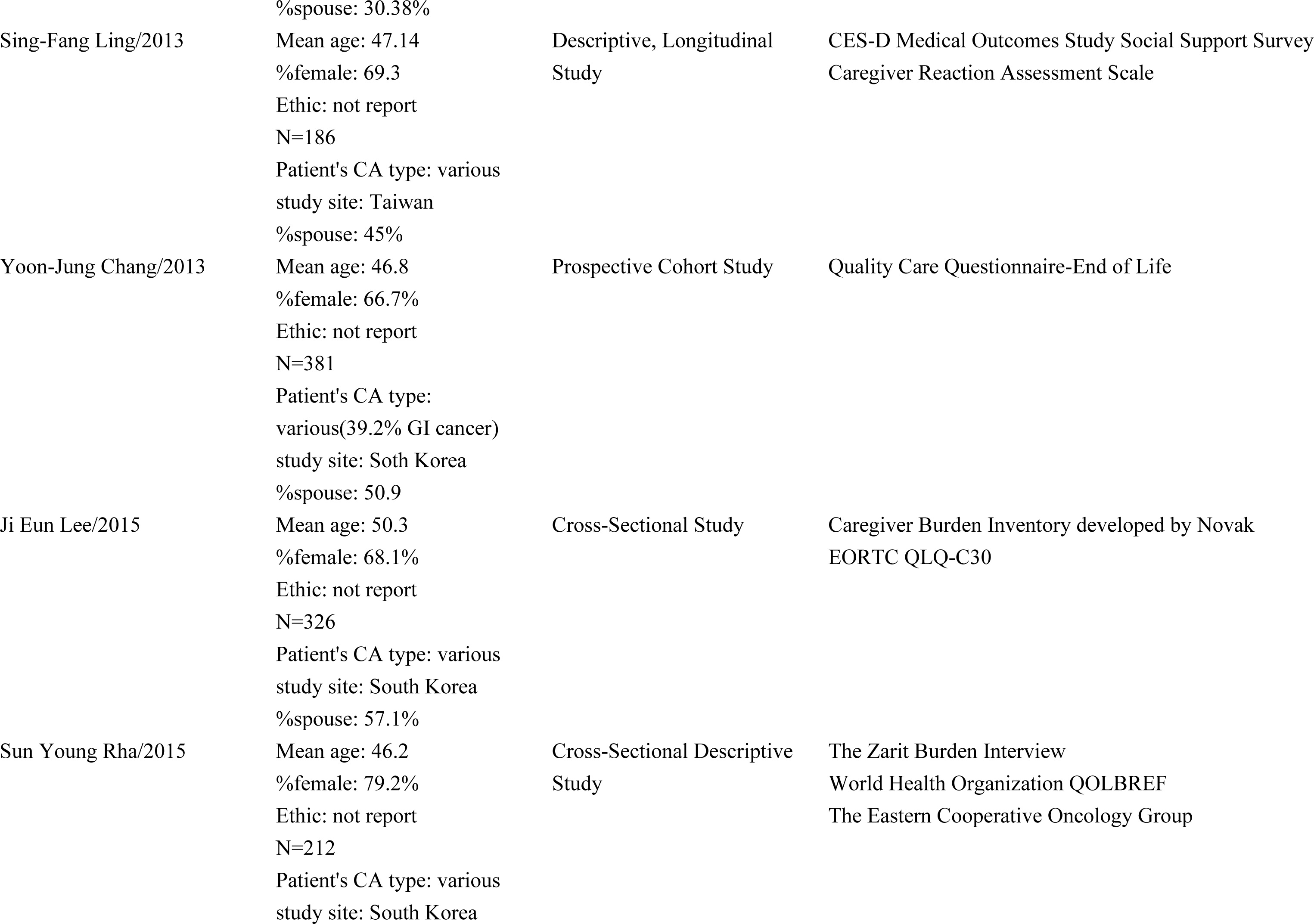
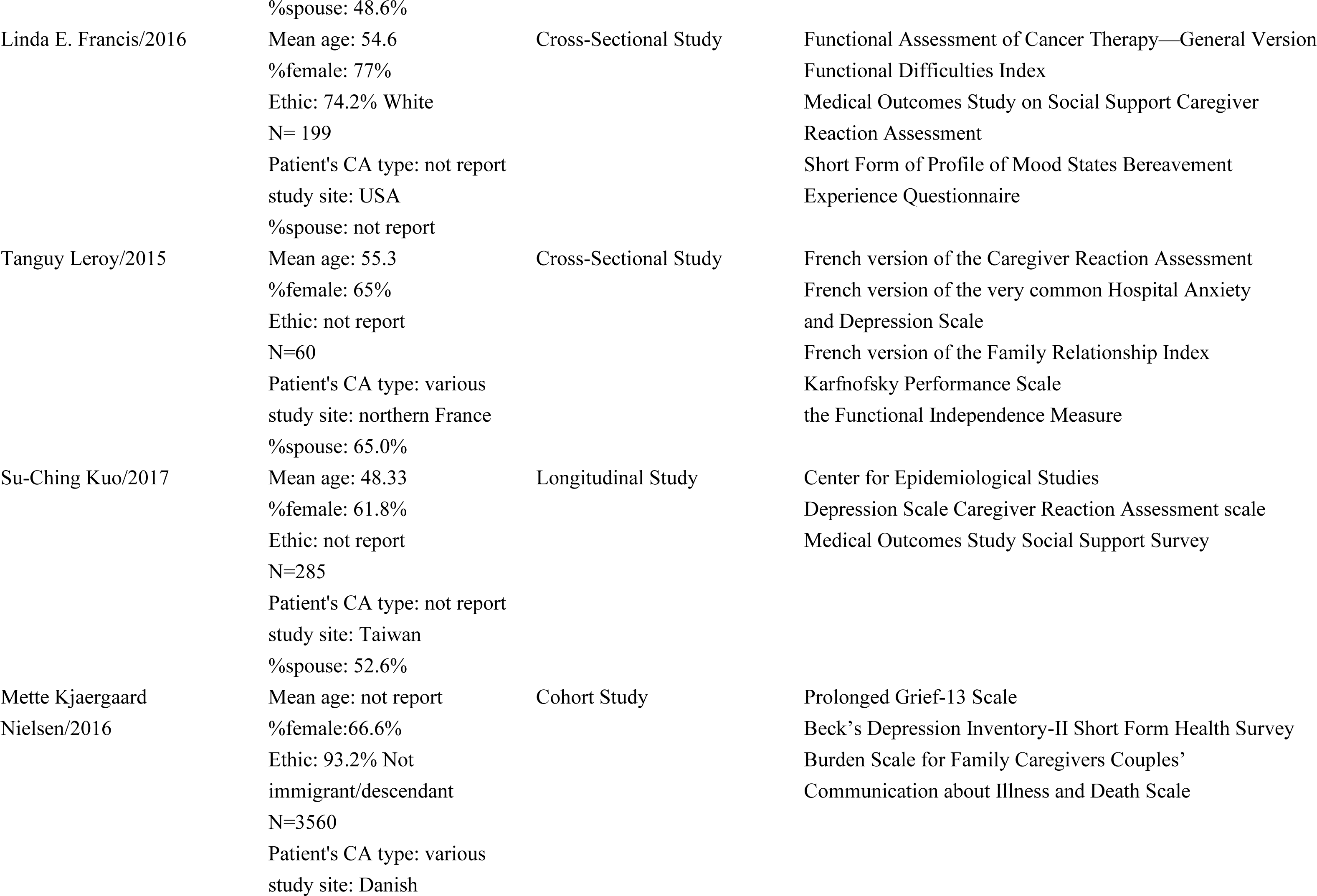
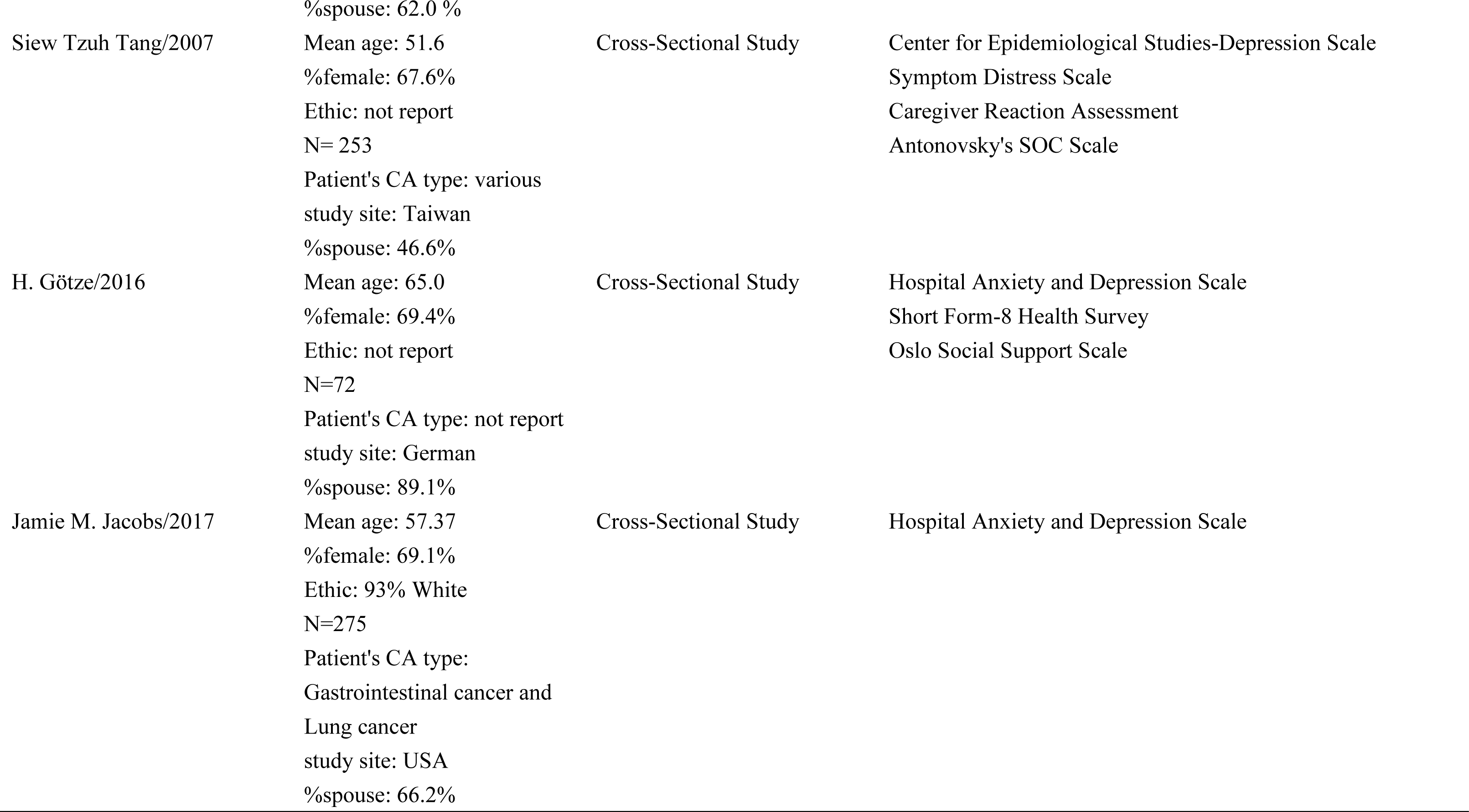
Descriptive studies evaluating psychosocial outcomes of cancer family caregivers during the palliative, hospice, and post-death bereavement phases.

**Table 2.**
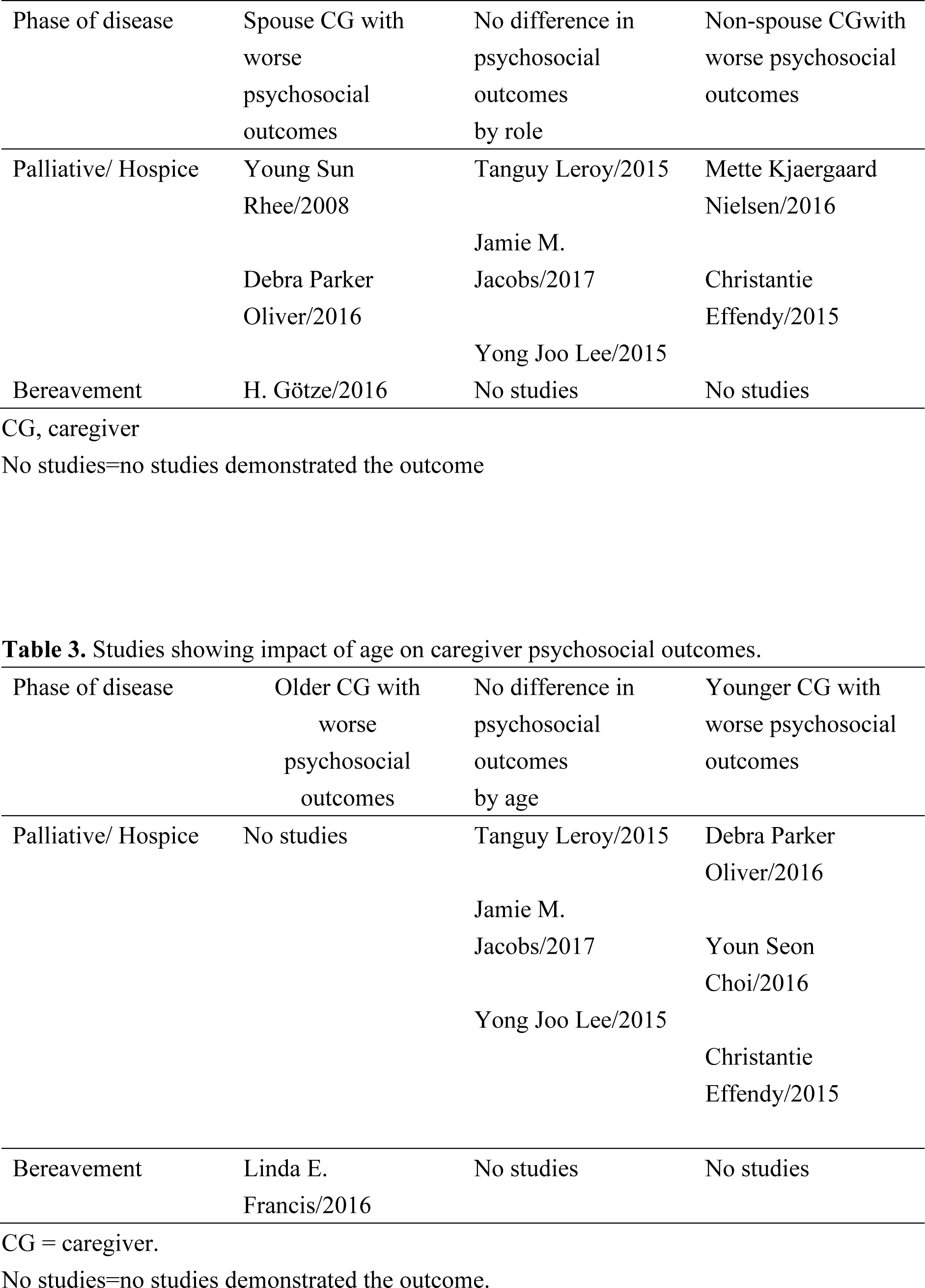
Studies showing impact of spouse versus non-spouse role on caregiver psychosocial outcomes.

Age was a predicted variable for provided inconsistent results frequently. Seven palliative studies examined age (Tanguy Leroy, 2015; Jamie M. Jacobs, 2017; Yng Joo Lee, 2015; Debra Parker Oliver, 2016; Youn Seon Choi, 2016; Christantie Effendy, 2015; Linda E. Francis, 2016). Three of the seven studies showed no influence from age (Tanguy Leroy, 2015; Jamie M. Jacobs, 2017; Yong Joo Lee, 2015), whereas three studies showed younger caregivers experiencing worse psychosocial outcomes than did older caregivers (Debra Parker Oliver, 2016; Youn Seon Choi, 2016; Christantie Effendy, 2015). One bereavement phase study viewed at age as a intermediate variable, and found that older age were relevant to depressed mood and grief (Linda E. Francis, 2016) (Table 3).

**Table 3.** Studies showing impact of age on caregiver psychosocial outcomes.

The effect of gender differences on psychosocial outcomes was reported with medicore, with six palliative/hospice phase studies (Young Sun Rhee, 2008; Beate Schrank, 2015; Tanguy Leroy, 2015; Jamie M. Jacobs, 2017; Christantie Effendy, 2015; Yong Joo Lee, 2015) and one bereavement phase studies (Linda E. Francis, 2015) examining the association. Most ordinarily, no significant influence from gender was found (Tanguy Leroy, 2015; Jamie M. Jacobs, 2017; Christantie Effendy, 2015; Yong Joo Lee, 2015). Two palliative/hospice phase studies (Young Sun Rhee, 2008; Beate Schrank, 2015) showed that female had worse psychosocial outcomes than male. One study also appointed out that female had worse psychosocial outcome during bereavement (Linda E. Francis/2015) (Table 4).

**Table 4.** Studies showing impact of gender on caregiver psychosocial outcomes.

| Phase of disease | Female CG with worse psychosocial outcomes | No difference in psychosocial outcomes by gender | Male CG with worse psychosocial outcomes |
| --- | --- | --- | --- |
| Palliative/ Hospice | Young Sun Rhee/2008<br>Beate Schrank/2015 | Tanguy Leroy/2015<br>Jamie M. Jacobs/2017<br>Christantie Effendy/2015<br>Yong Joo Lee/2015 | No studies |
| Bereavement | Linda E. Francis/2015 |  | No studies |

CG = caregiver.
No studies = no studies demonstrated the outcome.

For particular type of the demographic predictor variables of connection to patient, which was age and gender, the inconstant results across studies might be corresponded to whether the investigators controlled for confounders such as financial status, employment status, ethnicity, health status and any other factor that might occupy caregiver’s time and attention. While confounder was in control, differences by predictor variables were less likely to be identified.

Across the 23 studies examining family caregivers in the palliative, hospice, and bereavement phases, 53 unique instruments were used. There were several instruments being used; Medical Outcome Study Social Support Survey, Caregiver Reaction Assessment, Center for Epidemiological Studies-Depression scale, Hospital Anxiety and Depression Scale, Caregiver Quality of Life Index-Cancer scale appeared in nine times, six times, four times, four times and three times, respectively.

Although some instruments reused frequently, the vast majority of instruments used were with rare confirmation of the findings with an objective measure. As compare to MOS Social Support Survey of Chinese version (Reliability: all Alphas >0.93; Validity: 0.88∼0.99, 2004), it made barely differences in the original version (Reliability: all Alphas >0.91; Validity: 0.69∼0.82, 1991). So, we assumed that psychometric instrument would not be influenced by location, culture, language and time.

Coincidently, it was worth to mention that caregiver got worst status before patient’s death would positively be relevance to mechanisms active coping, feel burden, get level of daily life impairment; and negatively associated with strength of believe, social functioning, role emotional and mental health. After patient’s death, caregiver showed worst status on being staying mental health at baseline, mechanisms active coping, depressive symptom and quality of death assessed; and exacerbate in discrepancy between perceived and ideal levels of practical support, anxiety and depressive symptoms, role physical, general health, mental component summary, vitality, social functioning, role emotional and mental health. (Table 5) At last but not least, there were 16 factors, which was being younger, social support, environmental, patients with functional deterioration, adapting poorly, at the beginning of home care, caregiver’s perception of health, caring for a patient with poor ECOG, degree of kinship(partner/other), geographic location, hospitalized patients, poor health, shorter length of stay, the spouse of the patient, unable to function normally and younger age and poorer self-rated global health, were correlated with caregiver’s depression; 7 factors, also known as being younger, caregiver’s educational level, emotional distress, having no previous experience of caring, hospitalized patients, not being the spouse, physical function, were associated with caregiver’s quality of life; nine factors, which was being younger, social support, environmental, feeling burdened, prefer palliative care, symptoms of CMD, WHOQOL-BREF TOTAL, women, are relevant to caregiver’s burden; 7 factors, also known as being younger, environmental, caregiver’s perception of health, caring for a patient with poor ECOG, poorer self-rated global health, caregiver’s perception of money, grief, were concerned with caregiver’s anxiety (Table 6).

**Table 5.** Caregiver status before/after patient’s death.

|  | Caregiver worst status before<br>patient's death | Caregiver worst status after<br>patient's death |
| --- | --- | --- |
| Carer mental health at<br>baseline | NR | P |
| Coping mechanisms active<br>coping | P | P |
| Quality of death assessed by<br>carer | NR | N |
| Depressive symptoms | NR | P |
| Religion (strength of<br>believe) | N | NR |
| Coping mechanism<br>substance abuse | N | NR |
| Level of daily life<br>impairment | P | NR |
| QOL | N | NR |
| Burden | P | NR |
| Discrepancy between<br>perceived and ideal<br>levels of practical support | NR | N |
| Anxiety and depressive<br>symptoms | NR | P |
| Role physical | NR | N |
| General Health | NR | N |
| Mental component summary | N | N |
| Vitality | NR | N |
| Social functioning | N | N |
| Role emotional | N | N |
| Mental health | N | N |

P=Positive Correlation
N=Negative Correlation
NR=No Relevant

**Table 6.**
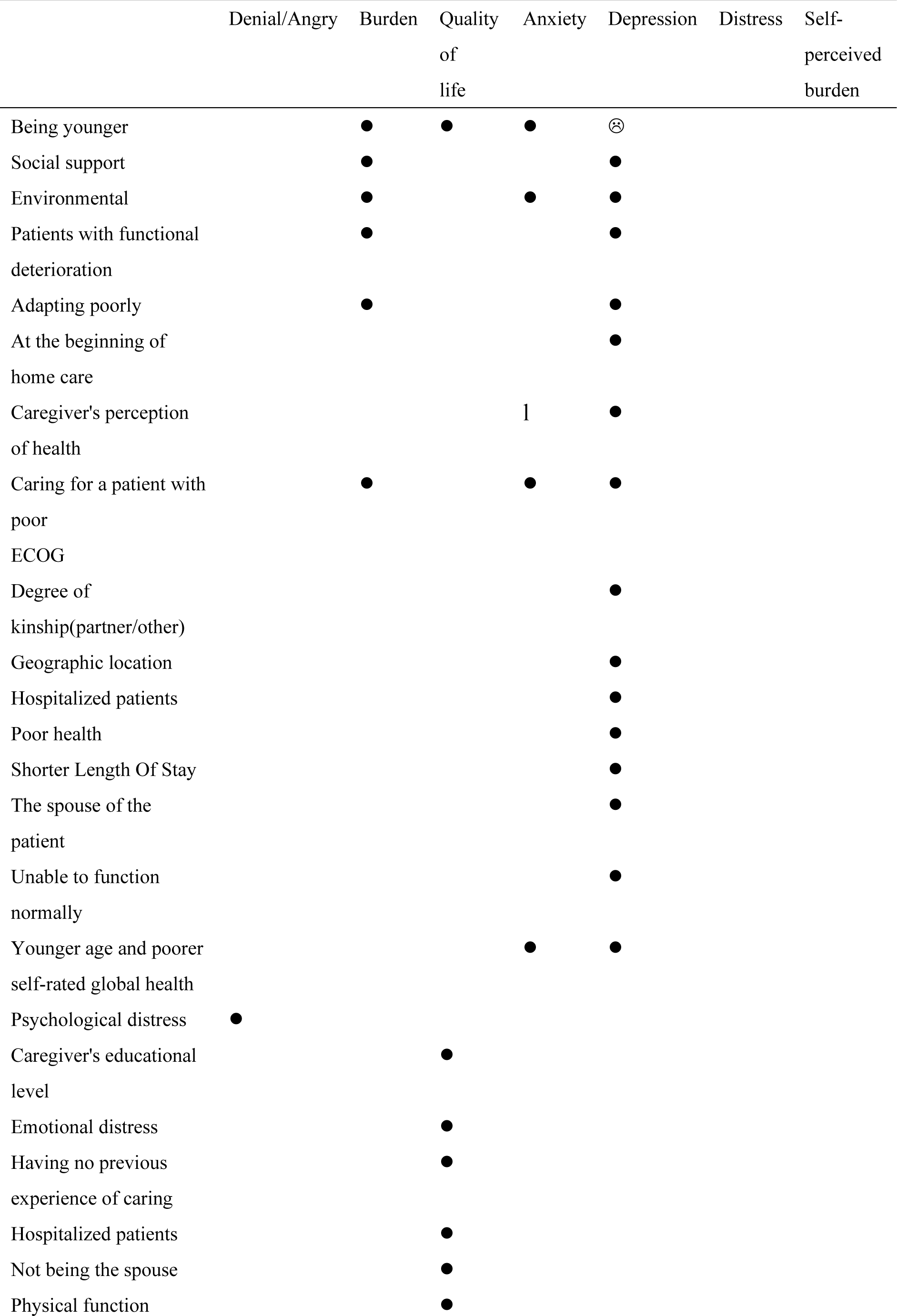
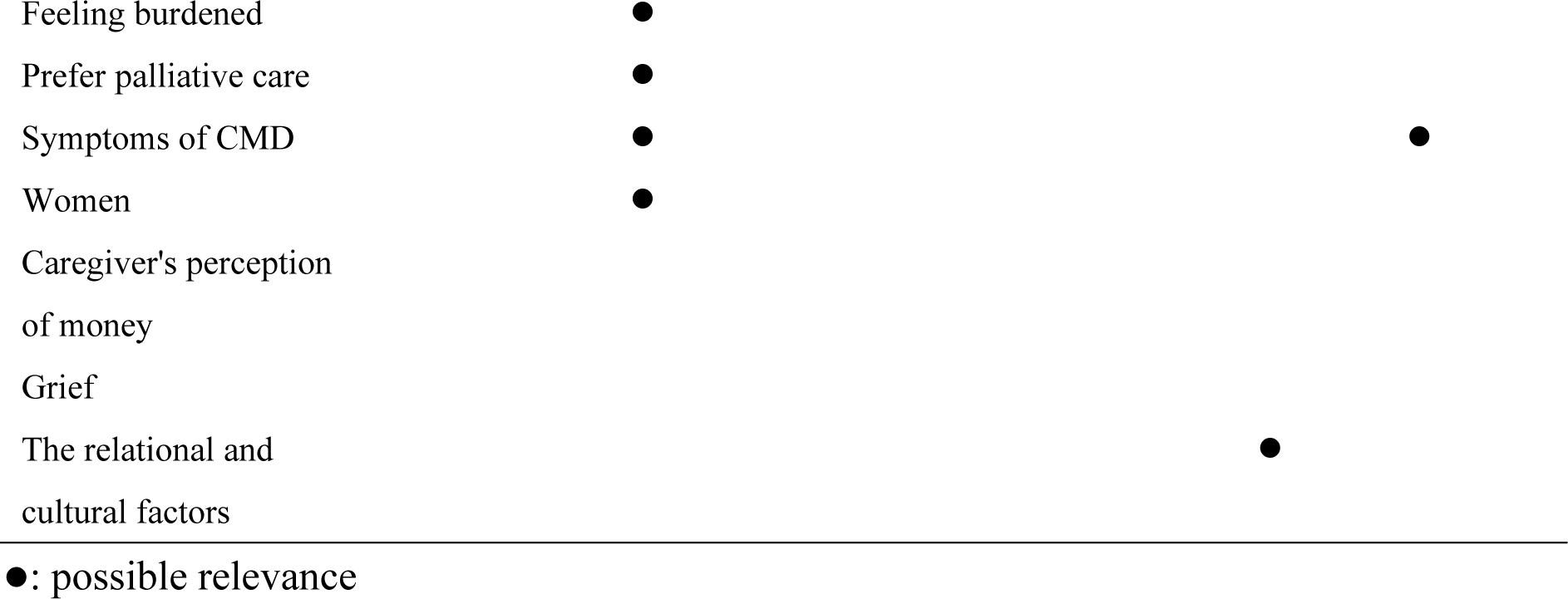
Influenced Factors \ Caregiver’s Perception.

## DISCUSSION

Descriptive systematic review of literature in family caregiver was difficult to handle and integrate, which research obstacle was as same as previous research (Annaleila Williams, 2010). The major of obstacle to aggregation was in poverty of research standard. Numerous unique measurement tools existed. All of studies were at high range of sample size and uses cross-sectional design thereby unable to provide information on the development of continuity and couldn’t show individual differences in development.

The definition of emotional express was staggering. Emotion, generally speaking, was a series of subjective experiences that produced a combination of feelings, thought and behavior (Denton, Derek, 2006). However, in regard to metal illness, researchers applied reliable measurement tool to illustrate potential sufferers’ status; it might be helpful for subjects to prevent their deterioration. Although original definition of emotion was divergent, result of reaction from caregivers during bereavement were with two theories, which was the relief model and the complicated grief model (Shear, M. K, 2011). Relief model (Bass, D. M., 1990) suggested that caregivers who suffered increasing strain would experience relief after the death of the care recipient. As the caregiving role ended, it was assumed that the caregiving strain vanishes at the same time. Furthermore, complicated grief model proposed that increasing caregiver role strain predicted poorer bereavement adjustment. It argued that caregiver stress escalated over time, leaving the individual with weaker coping resources to adjust to the bereavement experience. According to Table 5 and Table 6, it might be sorted out a possible outcome. If strength of believe and coping mechanism substance abuse were worthwhile, then family caregiver can make better quality of life. While taking care of patients, caregiver faced level of daily life impairment so that brought them burden. No matter what caregiver status before or after patient’s death, they were poor in mental health, social functioning and role emotional. Discrepancy between perceived and ideal levels of practical support and low score of mental component summary, it caused anxiety and depressive symptoms easily. Anxiety, depression and stress were influence by being younger, environment and low level of life quality; and low level of life quality was influenced by emotional distress and stress. It was mutually affected then lead family caregiver to worst situation. If quality of death assessed by carer did not achieve their satisfaction, while mental health was under baseline, depressive symptoms occur. After patient passed away, caregivers’ role physical and viability declined, which also meant deterioration of their general health. However, a subject of emotional reaction was worth to be viewed as personal difference, but it showed that the complicated grief model was suitable to our findings.

Many of variance, composed by personal status, hospitalized service and environmental factor, were highly frequently appeared in burden, quality of life, anxiety and depression. Comparatively, those variances were difficult to be exited independently; as a result, it was imperative to clarify their relationship also known as cause and effect. At the same time, 23 studies showed no culture difference in their attitude toward their loved one. As compared to previous research (Anna-Leila Williams, 2010), it showed same result on following, which cross section study design was more than longitude study design; sample in various cancer type was more than in specific type; spouses, gender, and age have no consistent influence on family caregivers; and number of studies on relationship between age and caregiver barely changed.

As previous research demonstrated that main study site was in the US, but, after time elapsed, Korea (7) was keeping pace with the US (7). However, we discovered that researcher used same material dividing into two pieces for studying; this phenomenon might derive from focusing on the number of papers and anxiously creating study research in short term. The influence of caregiver by spouses showed inconsistent result, but number of studies had been increased from three to nine. Unlike almost two decades ago, spouses were narrowly defined as relationship of bisexuality; it was assumed that people stood for their right, despite being mute, in the society. In contrast to previous study which reported 89 unique instruments including almost half of (n=41) instrument showing no psychometric testing; it was worthwhile to mention that there were 54 unique instruments containing less than a quarter (n=13) no psychometric study reported. As for instrument testing, MOS social support survey (Sherbourne, C. D., 1991), for example, started from definition of perception; hereafter, the selection of the pool of items was instructed by a strong a priori conceptual framework. Nineteen variables, selected from 50 items, were conducted in progress by multitrait and factor analyses in order to test their validity and reliability. The specificity and stability of psychometric instrument all went through rigorous study design, so that made results of the study being much more persuasive. As for the concerns in previous review, it had been changed, after suggestion, better than before.

## LIMITATION

Although it restricted from English, it might exclude other relevant research in different language. The major database was used in PubMed, but it used to be categorized as medicine and health, which, on the other words, humanities and social science would be out of option. Because the research after beginning of 2017 was excluded, the latest study may show different result which may led our inference indecisive.

## SUGGESTION

In spite of family caregivers paid their attention for their loved one with no regret, deterioration of health and financial crisis confronted against caregivers after time elapsed. It was a major issue to set them back in appropriate employment when they were out of society. Either opportunity of development or self-identity was precarious and in jeopardy, which exploited family caregiver’s psychosocial and physical wellbeing.

Despite of non-intervention study, if questionnaire can react the current status to cases, it was able to help them understanding their situation. The none-intervention study should belong to no intervene in the study; and researcher acted positively on supporting cases after questionnaire completed. While looking after of their loved one, family caregivers took patient as priority; in the meanwhile, erosion of carers’ self-identity had been virtually on the remaining life time of patient. With respect to regard caregiving as ethical responsibility, it was urgently considered caregiving, whether carer or cared one, as fundamental human rights; thus, caregiving should be a choice instead of obligation. Family caregivers were necessary to treat themselves well, seek for help, own their life, refuse being guilty, accept positive feedback and complete their career plan in the future.

## CONCLUSION

By integrating the descriptive study dedicates to carer during different phase, this literature review shoulders previous research. We showed same doubt as previous study. There was inconsistent result in spouse, gender, and age. However, it was much rigorous than before while using instrument, and also made changed on main study site and research on spouses. Afterwards, we inferred that the complicated grief was suitable to status of family caregiver after patient’s death. The physical and psychological issue of family caregiver in the palliative, hospice, and bereavement phases shall not be ignored; mostly, they are high-risk population.

## Data Availability

All data produced in the present work are contained in the manuscript

## REFERENCE

1. Francis, L. E., Kypriotakis, G., O’Toole, E. E., & Rose, J. H. (2016). Cancer patient age and family caregiver bereavement outcomes. Supportive Care in Cancer, 24(9), 3987–3996.

2. Chang, Y. J., Kwon, Y. C., Lee, W. J., Do, Y. R., Lee, K. S., Kim, H. T.,…& Yun, Y. H.. (2013). Burdens, needs and satisfaction of terminal cancer patients and their caregivers. Asian Pacific Journal of Cancer Prevention, 14(1), 209–215.

3. Lee, J. E., Shin, D. W., Cho, J., Yang, H. K., Kim, S. Y., Yoo, H. S.,…& Park, J. H. (2015). Caregiver burden, patients’ self-perceived burden, and preference for palliative care among cancer patients and caregivers. Psycho-Oncology, 24(11), 1545–1551.

4. Rha, S. Y., Park, Y., Song, S. K., Lee, C. E., & Lee, J. (2015). Caregiving burden and the quality of life of family caregivers of cancer patients: the relationship and correlates. European Journal of Oncology Nursing, 19(4), 376–382.

5. Götze, H., Brähler, E., Gansera, L., Schnabel, A., Gottschalk-Fleischer, A., & Köhler, N. (2018). Anxiety, depression and quality of life in family caregivers of palliative cancer patients during home care and after the patient’s death. European journal of cancer care, 27(2), e12606.

6. Rhee, Y. S., Yun, Y. H., Park, S., Shin, D. O., Lee, K. M., Yoo, H. J.,…& Kim, N. S. (2008). Depression in family caregivers of cancer patients: the feeling of burden as a predictor of depression. Journal of Clinical Oncology, 26(36), 5890–5895.

7. Francis, L. E., Kypriotakis, G., O’Toole, E. E., Bowman, K. F., & Rose, J. H. (2015). Grief and risk of depression in context: the emotional outcomes of bereaved cancer caregivers. OMEGA-Journal of Death and Dying, 70(4), 351–379.

8. Kuo, S. C., Chou, W. C., Chen, J. S., Chang, W. C., Chiang, M. C., Hou, M. M., & Tang, S. T. (2017). Longitudinal changes in and modifiable predictors of the prevalence of severe depressive symptoms for family caregivers of terminally ill cancer patients over the first two years of bereavement. Journal of palliative medicine, 20(1), 15–22.

9. Tang, S. T., & Li, C. Y. (2008). The important role of sense of coherence in relation to depressive symptoms for Taiwanese family caregivers of cancer patients at the end of life. Journal of Psychosomatic Research, 64(2), 195–203.

10. Parker Oliver, D., Washington, K., Smith, J., Uraizee, A., & Demiris, G. (2017). The prevalence and risks for depression and anxiety in hospice caregivers. Journal of palliative medicine, 20(4), 366–371.

11. Ling, S. F., Chen, M. L., Li, C. Y., Chang, W. C., Shen, W. C., & Tang, S. T. (2013, January). Trajectory and influencing factors of depressive symptoms in family caregivers before and after the death of terminally ill patients with cancer. In Oncology nursing forum (Vol. 40, No. 1).

12. Leroy, T., Fournier, E., Penel, N., & Christophe, V. (2016). Crossed views of burden and emotional distress of cancer patients and family caregivers during palliative care. Psycho-Oncology, 25(11), 1278–1285.

13. Jacobs, J. M., Shaffer, K. M., Nipp, R. D., Fishbein, J. N., MacDonald, J., El-Jawahri, A.,…& Greer, J. A. (2017). Distress is interdependent in patients and caregivers with newly diagnosed incurable cancers. Annals of Behavioral Medicine, 51(4), 519–531.

14. Nielsen, M. K., Neergaard, M. A., Jensen, A. B., Bro, F., & Guldin, M. B. (2016). Psychological distress, health, and socio-economic factors in caregivers of terminally ill patients: a nationwide population-based cohort study. Supportive Care in Cancer, 24(7), 3057–3067.

15. Zaider, T., & Kissane, D. (2009). The assessment and management of family distress during palliative care. Current opinion in supportive and palliative care, 3(1), 67.

16. Schrank, B., Ebert-Vogel, A., Amering, M., Masel, E. K., Neubauer, M., Watzke, H.,…& Schur, S. (2016). Gender differences in caregiver burden and its determinants in family members of terminally ill cancer patients. Psycho-Oncology, 25(7), 808–814.

17. Sutherland, N., Ward-Griffin, C., McWilliam, C., & Stajduhar, K. (2017). Structural impact on gendered expectations and exemptions for family caregivers in hospice palliative home care. Nursing inquiry, 24(1).

18. Choi, Y. S., Hwang, S. W., Hwang, I. C., Lee, Y. J., Kim, Y. S., Kim, H. M.,…& Koh, S. J. (2016). Factors associated with quality of life among family caregivers of terminally ill cancer patients. Psycho-Oncology, 25(2), 217–224.

19. Choi, Y. S., Hwang, S. W., Hwang, I. C., Lee, Y. J., Kim, Y. S., Kim, H. M.,…& Koh, S. J. (2016). Factors associated with quality of life among family caregivers of terminally ill cancer patients. Psycho-Oncology, 25(2), 217–224.

20. Effendy, C., Vernooij-Dassen, M., Setiyarini, S., Kristanti, M. S., Tejawinata, S., Vissers, K., & Engels, Y. (2015). Family caregivers’ involvement in caring for a hospitalized patient with cancer and their quality of life in a country with strong family bonds. Psycho-Oncology, 24(5), 585–591.

21. Wright, A. A., Keating, N. L., Balboni, T. A., Matulonis, U. A., Block, S. D., & Prigerson, H. G. (2010). Place of death: correlations with quality of life of patients with cancer and predictors of bereaved caregivers’ mental health. Journal of Clinical Oncology, 28(29), 4457.

22. Lee, Y. J., Kim, J. E., Choi, Y. S., Hwang, I. C., Hwang, S. W., Kim, Y. S.,…& Kim, S. J. (2016). Quality of life discordance between terminal cancer patients and family caregivers: a multicenter study. Supportive Care in Cancer, 24(7), 2853–2860.

23. Kim, S. H., Hwang, I. C., Ko, K. D., Kwon, Y. E., Ahn, H. Y., Cho, N. Y., & Kim, S. J. (2015). Association between the emotional status of family caregivers and length of stay in a palliative care unit: a retrospective study. Palliative & supportive care, 13(6), 1695–1700.

24. Kapari, M., Addington-Hall, J., & Hotopf, M. (2010). Risk factors for common mental disorder in caregiving and bereavement. Journal of pain and symptom management, 40(6), 844–856.

25. Colon, M. (2005). Hospice and Latinos: a review of the literature. Journal of social work in end-of-life & palliative care, 1(2), 27–43.

26. Sherbourne, C. D., & Stewart, A. L. (1991). The MOS social support survey. Social science & medicine, 32(6), 705–714.

